# Multimodal Freezing of Gait Detection: Analyzing the Benefits of Physiological Data

**DOI:** 10.1101/2024.10.25.24315880

**Authors:** Po-Kai Yang, Benjamin Filtjens, Pieter Ginis, Maaike Goris, Alice Nieuwboer, Moran Gilat, Peter Slaets, Bart Vanrumste

## Abstract

Freezing of gait (FOG) is a debilitating symptom of Parkinson’s disease (PD), characterized by an absence or reduction in forward movement of the legs despite the intention to walk. Detecting FOG during free-living conditions presents significant challenges, particularly when using only inertial measurement unit (IMU) data, as it must be distinguished from voluntary stopping events that also feature reduced forward movement. Influences from stress and anxiety, measurable through galvanic skin response (GSR) and electrocardiogram (ECG), may assist in distinguishing FOG from normal gait and stopping. However, no study has investigated the fusion of IMU, GSR, and ECG for FOG detection. Therefore, this study introduced two methods: a twostep approach that first identified reduced forward movement segments using a Transformer-based model with IMU data, followed by an XGBoost model classifying these segments as FOG or stopping using IMU, GSR, and ECG features; and an end-to-end approach employing a multi-stage temporal convolutional network to directly classify FOG and stopping segments from IMU, GSR, and ECG data. Results showed that the two-step approach with all data modalities achieved an average F1 score of 0.728 and F1@50 of 0.725, while the end-to-end approach scored 0.771 and 0.759, respectively. However, no significant difference was found compared to using only IMU data in both approaches (p-values: 0.466 to 0.887). In conclusion, adding physiological data does not provide a statistically significant benefit in distinguishing between FOG and stopping.

## I. Introduction

**P**ARKINSON’S disease (PD) affects over six million people globally. Many PD patients experience a symptom called freezing of gait (FOG), which is defined as a sudden inability to move forward despite the intention to walk, posing risks of falls and reduced quality of life [1]–[3]. Although commonly referred to as a singular phenomenon, FOG is notably heterogeneous, manifesting in various forms: 1) rapid shuffling characterized by very short steps and insufficient foot clearance; 2) trembling in place, marked by alternating tremulous leg oscillations with minimal or no forward progression; and 3) pure akinesia, where there is little to no visible movement in the lower limbs [4]. Current FOG treatments primarily involve dopaminergic medications and physical therapy, adjusted based on FOG severity, which is typically assessed using subjective measures such as the New Freezing of Gait Questionnaire (NFOGQ) [5]. These tools, however, are prone to recall bias and lack sensitivity [6]. Semi-objective evaluation of FOG severity typically involves standardized tasks such as the Timed-Up and Go (TUG) [7] and 360 degree turning tasks [8] in clinical centers, with post-hoc visual analysis of video recordings serving as the gold standard for FOG assessment [9]. Nonetheless, this manual annotation process is labor-intensive and time-consuming, prompting the exploration of automatic approaches leveraging machine learning (ML) and deep learning (DL) models [10]–[13].

Monitoring FOG severity in daily life requires at-home solutions. Video recordings, while common, raise privacy concerns and are limited by the camera’s field of view [14]– [16]. Wearable sensors, such as inertial measurement units (IMU), offer a privacy-respecting alternative but require extensive training on diverse datasets to ensure reliable accuracy [17]. However, data collected in controlled laboratory settings often lacks the complexity of real-life scenarios, such as the variability in gait and volitional stopping periods [18]. These stopping periods, characterized by the absence of lower limb movements, pose challenges in differentiating from akinetic freezing [19]. Recent research has incorporated these stopping periods into training regimes to enhance detection performance in realistic conditions [12], [18], [20], [21]. Although such adaptations improve detection accuracy [12], relying solely on IMU signals may be insufficient for distinguishing between volitional stopping and FOG [19], [22].

Factors such as stress and anxiety are recognized contributors to the occurrence of FOG [23], [24], influencing physiological signals such as electrocardiogram (ECG) and galvanic skin response (GSR), which are indicators of emotional states [25], [26]. ECG signals have been investigated for their potential to enhance FOG detection, as heart rate (HR) and heart rate variability increase during FOG-episodes, albeit not consistently [19], [22], [27], [28]. However, since these metrics also rise during physical activity including gait [19], [28] and are highly influenced by cognitive load [23], [29], their specificity for detecting FOG is limited [28]. Conversely, increases in GSR-related features occur just before and after FOG episodes, distinguishing FOG from other gait activities [28]. Until now, most studies have used GSR and ECG features tailored for statistical testing rather than for ML-based model classification. For example, Cockx *et al*. imposed strict criteria on FOG trials, excluding FOG events, stopping, or normal gait events preceded by another FOG episode within six seconds [19]. Maiden *et al*. included only FOG episodes that occurred during turns in their analysis [27]. These approaches may not reflect everyday scenarios where FOG and stopping frequently occur in close succession, and FOG can occur during more than just turning. Moreover, while HR alone may not reliably differentiate FOG from normal gait, integrating it with IMU and GSR signals could potentially improve accuracy. Thus, Cockx *et al*. suggested using IMU data to identify movement reduction first and then utilize ECG characteristics to determine whether the reduction is due to FOG or stopping, yet this approach still requires validation [19]. Additionally, when monitoring FOG severity in daily life, patients’ movements are often accompanied by other tasks that place a high cognitive load, such as walking through doorways or when performing a concurrent dual-task (DT) when walking [30]–[34]. These cognitively demanding tasks could potentially influence the patients’ stress and anxiety levels [35], [36], which, in turn, may affect the utility of physiological data such as HR in distinguishing FOG from volitional stopping [19]. However, previous FOG detection studies [22], [28], [37], [38] have not thoroughly evaluated the impact of DT on the effectiveness of FOG detection under such conditions.

To the best of our knowledge, this study is the first to investigate the benefits of integrating motor (IMU) and physiological (GSR, ECG) data for FOG and stopping event detection using ML and DL. Unlike previous studies designed to investigate the mechanisms of GSR and ECG on FOG [19], [28], our study aims to clarify their added value in FOG detection during typical scenarios, such as TUG tests that include stopping periods and cognitive load DTs, which better simulate at-home conditions. This study hypothesized that adding physiological data would enhance detection and tested this with two approaches. First, we developed a two-step approach, building on Cockx *et al*. [19], which employs a traditional white-box ML classifier to categorize segments of reduced forward progression as FOG or stopping based on multimodal features. We identified these segments either manually, to evaluate the contribution of each modality, or automatically, using a DL-based segmentation model. Second, we introduced an end-to-end approach that directly detects FOG and stopping from raw multimodal signals using the segmentation model.

## II. Related work

### A. FOG detection based on IMU signals

Historically, FOG detection primarily utilized manual feature engineering with IMUs. Early methods used threshold algorithms such as the Freezing Index, which was defined as the ratio of power in the “freezing” band (3-8 Hz) to that in the “locomotor” band (0.5–3 Hz), to differentiate FOG from non-FOG episodes [39]. Enhancements included integrating stride features, energy thresholds, and turn counts [40]–[42]. Advances followed with ML models such as decision trees and AdaBoost, applying hand-engineered IMU features [43]. Further developments by Shi *et al*. combined these features with wavelet analysis, creating 67 features that demonstrated superior performance in FOG detection on a dataset of 63 PD patients, with 486 FOG episodes using eXtreme Gradient Boosting (XGBoost) [13].

The diversity of FOG presentations has driven the shift towards end-to-end DL models that process raw data directly, avoiding manual feature engineering. Notable developments include Zhang *et al*.’s DeepCNN-LSTM model trained on IMU spectrograms [44] and Li *et al*.’s combination of a Temporal Convolutional Network (TCN) with LSTM for enhanced detection [45]. Enhancing these techniques, Shi *et al*. processed IMU data using continuous wavelet transform and utilized convolutional neural networks for FOG detection [13]. Building on these advancements, our recent study used a TCN for sample-level FOG annotation, with a subsequent prediction refinement block using a multi-stage TCN (MS-TCN) to reduce oversegmentation errors, facilitating the detection of the onset and offset of FOG episodes [12].

### B. FOG detection based on GSR and ECG signals

Research on using solely GSR or ECG signals for automatic FOG detection is limited. Mazilu *et al*. developed a GSR-based model using multivariate Gaussian distribution that predicted 71.3% of FOG episodes on a dataset of 11 subjects (#FOG = 184) by analyzing specific changes in skin conductance (SC) [28]. However, their ECG-based model struggled with reliability due to noise interference from body movements during walking.

The potential of multimodal data to enhance FOG detection was utilized by Mesin *et al*., who combined accelerometer (ACC), GSR, EMG, and EEG data, finding that performance dropped slightly when using a single sensor compared to multimodal setups [37]. They conducted the experiments on a dataset of 12 subjects (#FOG = 334). Zhang *et al*. extended this by testing combinations of EEG, EMG, ACC, and GSR features, showing that multimodal models significantly outperformed single modality models, achieving F1 scores above 90% on the same dataset [38]. Additionally, their findings indicated that models trained with solely GSR features had slightly higher F1 scores than those trained with ACC features alone, while a combination of features resulted in a significantly improved F1 score.

## III. Methods

This study explored the added value of combining physiological data (GSR, ECG) with motor data (IMU) for FOG detection. We introduced two approaches: a two-step approach, where an action segmentation model forms as a segmentation block detects forward movement reduction segments followed by a differentiation block classifying these as FOG or stopping (Figure 1a); and an end-to-end approach, where the model directly detects FOG and stopping segments (Figure 1b).

**Fig. 1.**
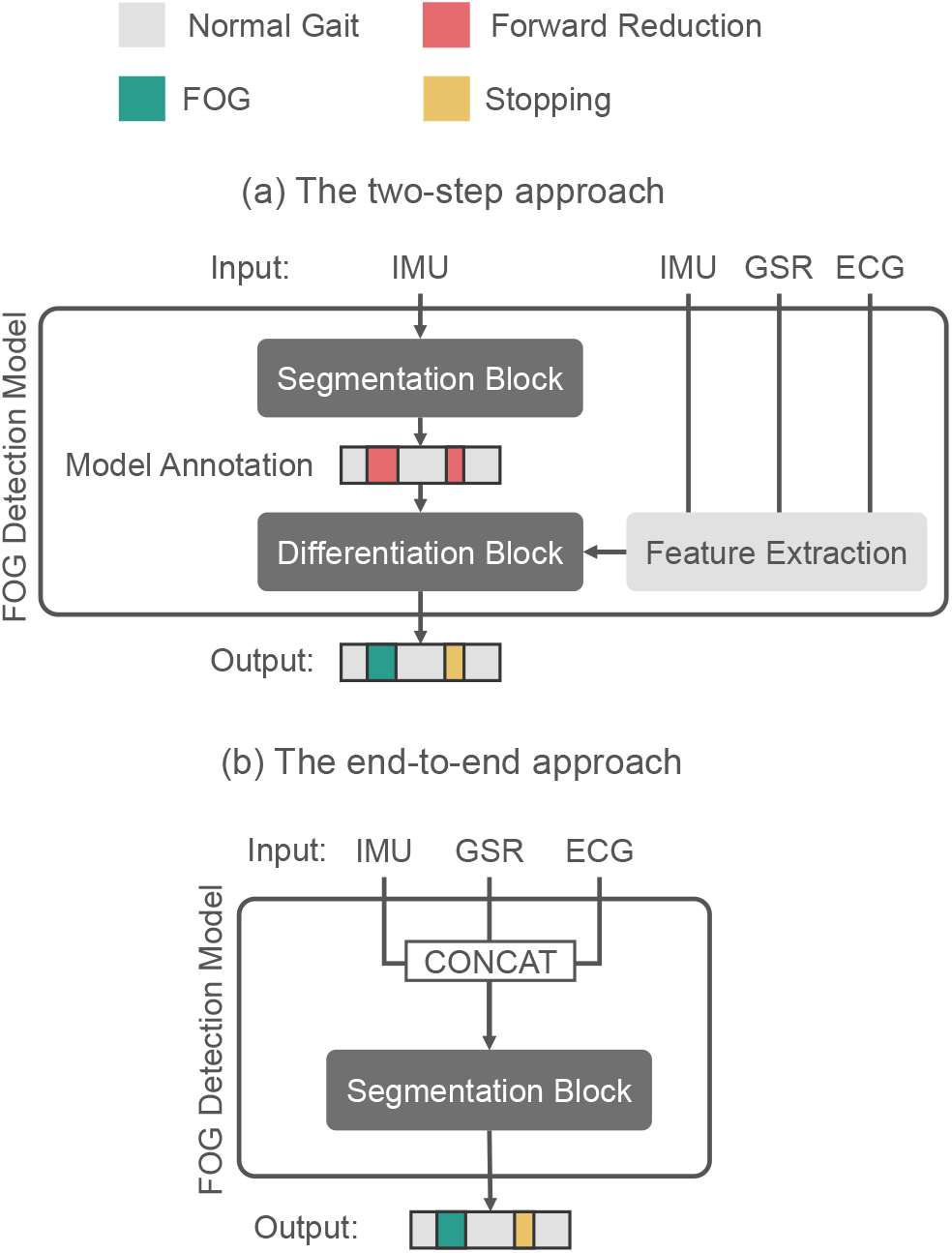
An overview of our two-step and end-to-end approach: (a) Two-step approach: The segmentation block identifies forward movement reduction segments, which are then classified as FOG or stopping by the differentiation block. (b) End-to-end approach: The segmentation block directly classifies segments as FOG or stopping.

We conducted three experiments: (1) model selection for the blocks within each approach, (2) evaluation of the best-performing model for the differentiation block using various modality combinations (IMU, GSR, ECG; IMU+GSR, IMU+ECG, GSR+ECG; IMU+GSR+ECG) and an analysis of the contribution of individual features, and (3) a comparison of the two approaches using IMU data alone versus combined IMU, GSR, and ECG data.

### A. Problem definition

An IMU trial can be represented as 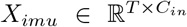, where *T* is the number of samples, and *C*_*in*_ is the input feature dimension. For IMU signals, *C*_*in*_ equals to 12 for acceleration and gyroscope data from two IMUs. Each IMU trial is associated with a GSR signal, which can be represented as 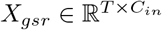, where *C*_*in*_ = 2 for C0 and C1. Additionally, the IMU and GSR trial is associated with an ECG trial 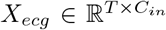, where *C*_*in*_ = 1 and *T*. HR extracted from ECG is denoted as 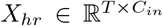, where *C*_*in*_ = 1. Each input trial *X*_*imu*_, *X*_*gsr*_, *X*_*ecg*_, and *X*_*hr*_ is associated with a ground truth label vector *Y* ^*T ×L*^, where the label *L* represents the manual annotation of FOG by the clinical experts.

#### 1) The two-step approach

First, an action segmentation model identifies segments with reduced forward movement. These segments are then classified by a traditional ML model to differentiate between FOG and stopping.

In this approach, the IMU data is processed by the segmentation model to produce prediction of shape 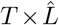, where 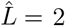 (0 for non-forward reduction and 1 for forward reduction segments, including FOG and stopping). Features from IMU, GSR, and ECG data are extracted for each predicted segment. The differentiation block subsequently classifies these forward reduction segments as either FOG or stopping events. To train the differentiation block, segments manually annotated by experts were used (as shown in Figure5), while during inference, segments identified by the segmentation block were employed (as shown in Figure1a). To determine the best model for the differentiation block, we compared four ML models: XGBoost [46], Support Vector Machine (SVM), K-Nearest Neighbors (KNN), and Decision Tree. For the segmentation block, we evaluated four action segmentation models: Bi-LSTM [47], Dilated TCN [48], MS-TCN [49], and ASFormer [50].

#### 2) The end-to-end approach

The segmentation model directly classifies segments into FOG, stopping, or non-forward reduction movements, bypassing the intermediate segmentation step. This aligns with recent studies that treat FOG detection as an action segmentation task [11], [12].

In the end-to-end approach (as shown in Figure1b), IMU, GSR, and HR data are combined along the feature dimension into a *T ×* 15 input (with HR replacing ECG for aligned sampling rates). This input is processed by an action segmentation model, producing 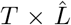 annotations, where 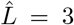 (0 for non-forward reduction, 1 for FOG, 2 for stopping). The best-performing model among Bi-LSTM, Dilated TCN, MS-TCN, and ASFormer was used for this approach.

### B. Data collection

This study included 18 PD patients (4 female, 14 male) with FOG, each with at least one self-reported daily FOG episode lasting a minimum of five seconds in the previous month. All subjects provided informed consent, and the study was approved by the Ethics Committee Research UZ/KU Leuven, with protocol number S65059. The average age was 67.33 *±* 6.71 years with an average disease duration of 12.39 *±* 5.01 years. Participant characteristics are detailed in Table I. The IMU portion of the dataset was previously utilized in our other studies [12], [51].

**TABLE I.**
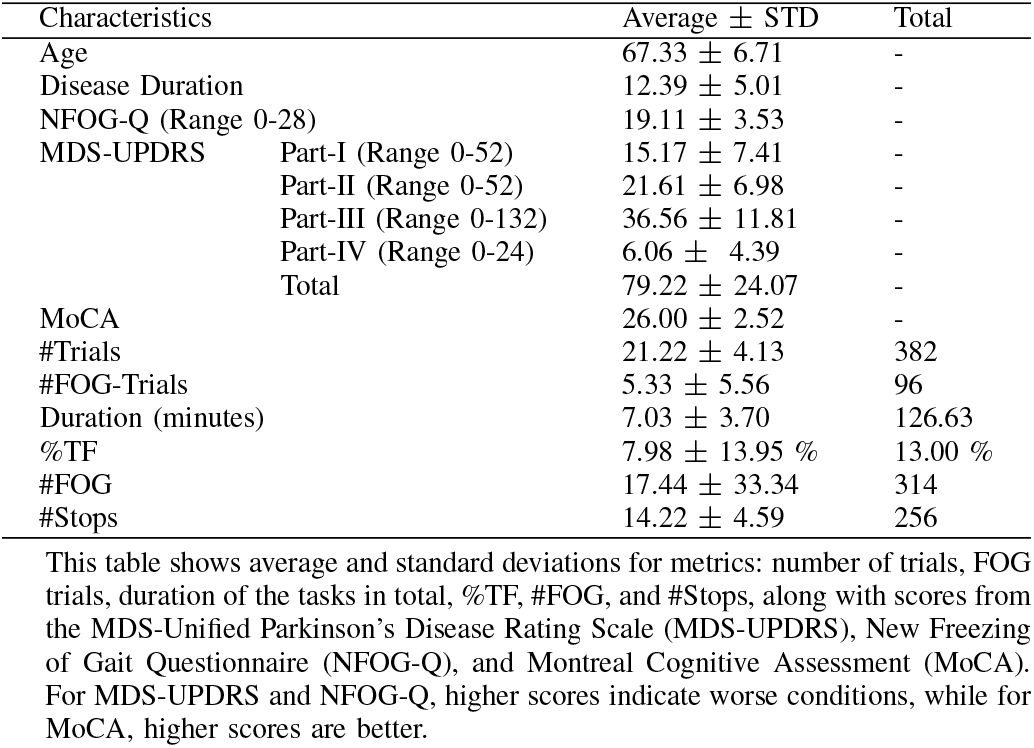
Participant and data characteristics

| Characteristics | Average ± STD | Total |
| --- | --- | --- |
| Age | 67.33 ± 6.71 | - |
| Disease Duration | 12.39 ± 5.01 | - |
| NFOG-Q (Range 0-28) | 19.11 ± 3.53 | - |
| MDS-UPDRS |  |  |
| Part-I (Range 0-52) | 15.17 ± 7.41 | - |
| Part-II (Range 0-52) | 21.61 ± 6.98 | - |
| Part-III (Range 0-132) | 36.56 ± 11.81 | - |
| Part-IV (Range 0-24) | 6.06 ± 4.39 | - |
| Total | 79.22 ± 24.07 | - |
| MoCA | 26.00 ± 2.52 | - |
| #Trials | 21.22 ± 4.13 | 382 |
| #FOG-Trials | 5.33 ± 5.56 | 96 |
| Duration (minutes) | 7.03 ± 3.70 | 126.63 |
| %TF | 7.98 ± 13.95 % | 13.00 % |
| #FOG | 17.44 ± 33.34 | 314 |
| #Stops | 14.22 ± 4.59 | 256 |

This table shows average and standard deviations for metrics: number of trials, FOG trials, duration of the tasks in total, %TF, #FOG, and #Stops, along with scores from the MDS-Unified Parkinson's Disease Rating Scale (MDS-UPDRS), New Freezing of Gait Questionnaire (NFOG-Q), and Montreal Cognitive Assessment (MoCA). For MDS-UPDRS and NFOG-Q, higher scores indicate worse conditions, while for MoCA, higher scores are better.

Participants underwent TUG tests to provoke FOG. They were instructed to rise from a chair, walk 2.5 meters, turn, return, and sit. Tests were conducted under both OFF-medication (12 hours after the last dose) and ON-medication (one hour post dose) states, both with and without self-generated or researcher-imposed stopping. Additionally, they performed the normal tasks (i.e., without stopping) with a DT, the auditory Stroop task [52], with the order of testing randomized to control for fatigue and learning. Each subject completed 24 TUG trials: two normal, two with DT, four normal with stopping in the turn, and four normal with stopping in a straight line, totaling 12 trials OFF and 12 trials ON medication.

Data collection utilized Shimmer3 IMU sensors (64 Hz) positioned on top of both feet, alongside a Shimmer3 GSR+ sensor (64 Hz) monitoring GSR. ECG sensors (512 Hz) were placed at torso positions: the left arm (LA), right arm (RA), left leg (LL), and right leg (RL). All sensor placements (Figure 2) and settings followed the manual of Shimmer [53]. RGB videos were recorded at 30 frames-per-second for offline FOG annotation performed iteratively by two expert raters using the Elan software [9]. FOG episodes were defined as periods where no effective forward steps could be made, starting with an ineffective step and ending after at least two effective steps, which were not included in the episode duration [1], [9].

**Fig. 2.**
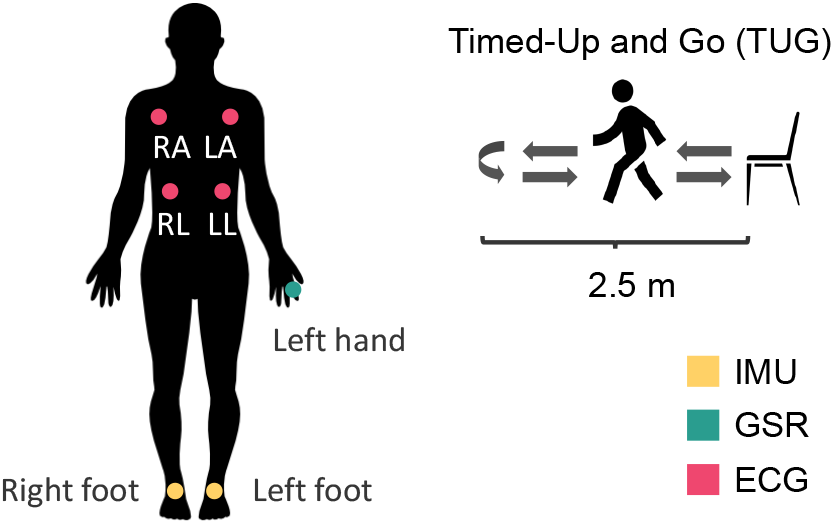
An illustration of sensor positions and TUG test protocol.

The dataset included 382 trials, totaling 126.63 minutes with 13.00% of the time annotated as FOG. It contained 314 FOG episodes (204 in single task (ST) trials, 110 in DT trials) and 256 stopping episodes (255 in ST trials, 1 in DT trials). Fifteen of the eighteen participants experiencing FOG, averaging 17.44 *±* 33.34 episodes each (median = 4). Details are presented in Table I.

### C. Data preprocessing

IMU signals were mean-centered to remove bias [12]. GSR signals were filtered with a third-order low-pass filter at 0.9 Hz, producing the filtered signal (C0) and its first derivative (C1) [28], [54], [55]. Following the procedure in [19], ECG signals were processed by selecting the highest-quality lead (i.e., lead II), applying a fifth-order bandpass filter (1–40 Hz), and detecting R-peaks using the Pan Tompkins algorithm [56] implemented via NeuroKit2 [57]. The average ECG quality score was 0.743 [57], with 86% of the signals rated as excellent [58]. Based on these metrics, we considered the ECG quality in our dataset suitable for further analysis [59]. HR was calculated using NeuroKit2 [57] from ECG and resampled to 64 Hz to synchronize with IMU and GSR signals. Figure 3 shows an example of synchronized IMU, C0, C1, ECG, and HR data.

**Fig. 3.**
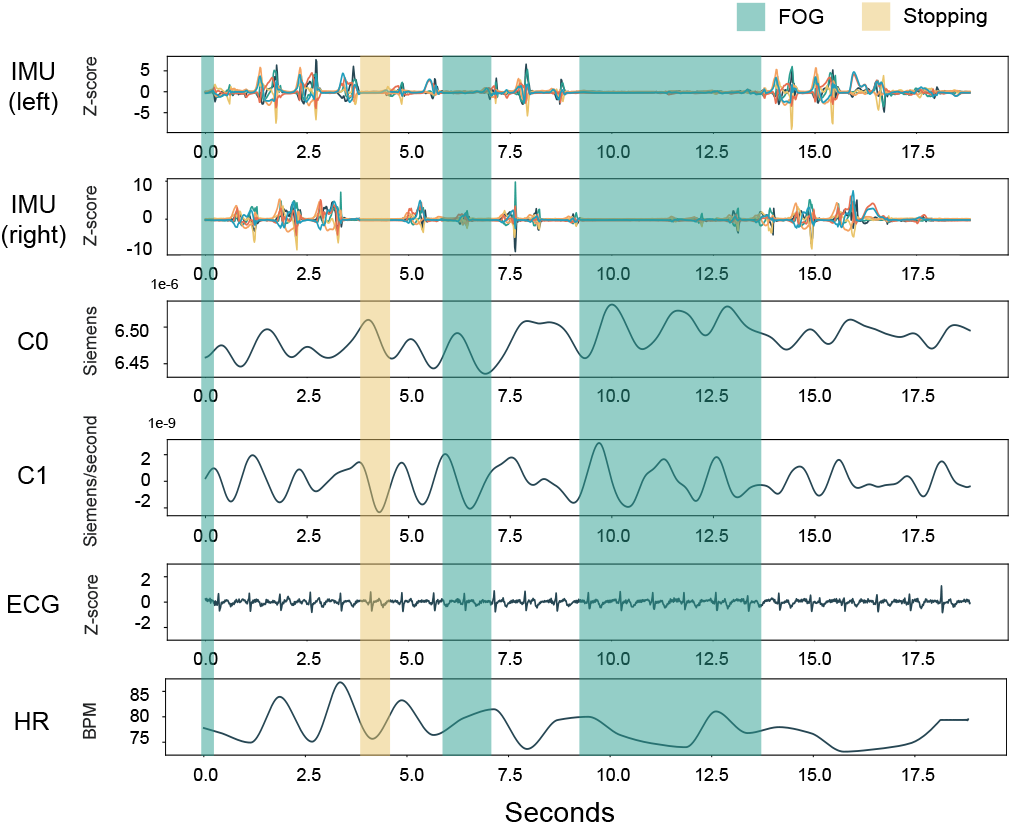
An illustration of the processed IMU, GSR (C0 and C1), ECG, and HR. IMU signals were z-normalized for better visualization. Experts’ annotations of FOG and stopping episodes are highlighted in green and yellow, respectively.

#### Feature generation

For the differentiation block, each FOG and stopping segment was associated with three time windows: pre-event (3 seconds before), event (the segment itself), and post-event (3 seconds after). The 3-second window size, chosen to capture an average of three heartbeats, balances accurate feature extraction with minimal latency [19], [28], [38]. From these windows, 18 features listed in Table II were extracted per window. Additionally, features were derived from the differences between the pre-event and event windows, and between the event and post-event windows, leading to a total of 90 features per segment, as shown in Figure 4. These features were used to classify each segment in the differentiation block.

**Fig. 4.**
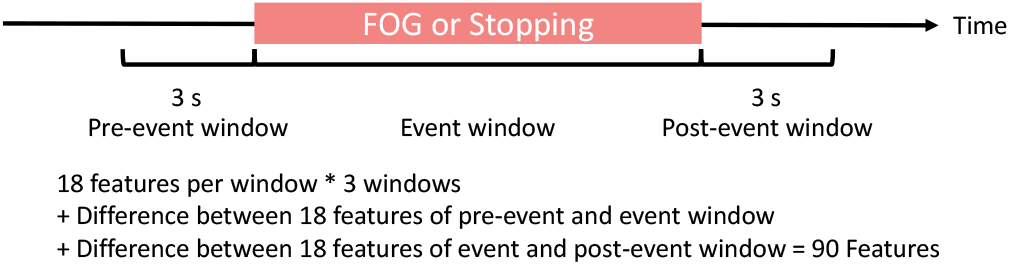
An illustration of feature generation for FOG and stopping segments: features are calculated within three windows: 3s before the event (pre-event), during the event, and 3s after the event (post-event).

**TABLE II.**
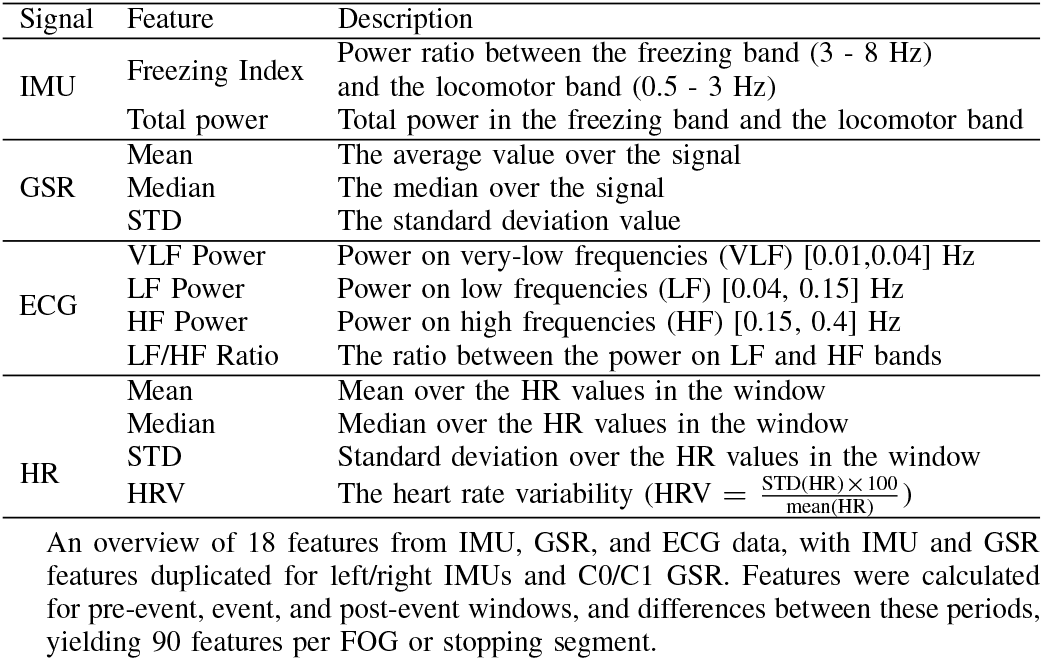
Features extracted for each windows

### D. Experimental setting

The experimental settings and metrics used in the three experiments are detailed in the following subsections.

#### 1) Experiment 1 (Model selection)

We used leave-one-subject-out cross-validation (LOSO-CV) to compare four ML models for the differentiation block. Each subject was tested individually, with the remaining subjects used for training and hyperparameter tuning. Validation data consisted of 20% of the training set, randomly selected from 4 of 17 subjects. To ensure that evaluation was not influenced by the segmentation block’s performance, the differentiation block was fed with experts’ manual annotations (as shown in Figure 5). For the differentation block, the hyperparameters for XGBoost, SVM, KNN, and Decision Tree models were tuned, with Table III listing the specific parameters for each model. Optimal settings were selected based on validation performance. For the segmentation block, Bi-LSTM, Dilated TCN, MS-TCN, ASFormer, were evaluated using LOSO-CV, with hyperparameters based on original studies [47]–[50].

**Fig. 5.**
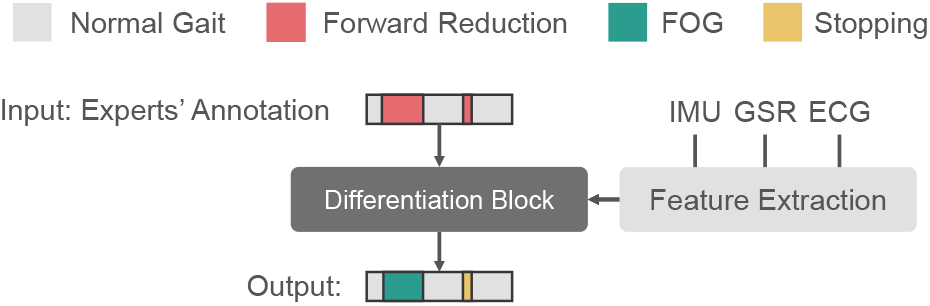
An illustration of the evaluation process for the differentiation block. To ensure an unbiased comparison of models and assess feature importance, expert-annotated segments are used as input during the evaluation of the differentiation block.

**TABLE III.** Hyperpamareter tuned for the ML models

| ML Model | Hyperparameter | Value |
| --- | --- | --- |
| XGBoost | max_depth | 1, 3, 5, 7, 9 |
|  | learning_rate | 0.1, 0.01, 0.001 |
|  | subsample | 0.5, 0.7, 1.0 |
|  | min_child_weight | 1, 3, 5, 7, 9 |
|  | n_estimators | 1, 3, 5, 7, 9 |
| SVM | C | 0.1, 1, 10, 100, 1000 |
|  | gamma | 1, 0.1, 0.01, 0.001, 0.0001 |
|  | kernel | 'rbf', 'linear' |
|  | class_weight | 'balanced', None |
| KNN | n_neighbors | 1, 2, 3, 4, 5, 6, 7, 8, 9, 10 |
|  | leaf_size | 20, 25, 30, 35, 40 |
|  | p | 1, 2 |
|  | weights | 'uniform', 'distance' |
|  | metric | 'minkowski', 'chebyshev' |
| Decision Tree | max_features | 'auto', 'sqrt', 'log2' |
|  | ccp_alpha | 0.1, 0.01, 0.001 |
|  | max_depth | 5, 6, 7, 8, 9 |
|  | criterion | 'gini', 'entropy' |

This table presents the hyperparameters tuned for the ML models. The optimal settings were determined based on validation performance using the LOSO-CV approach.

To evaluate the differentiation block, AUROC was computed across 570 segments. Sample-wise F1 scores were assessed per trial, classifying each sample as True Positive (TP), False Positive (FP), or False Negative (FN). Segmentwise F1@50 score was used to address over- and under-segmentation, where a segment is TP if its intersection over union with a ground-truth segment exceeds 0.5; otherwise, it is FP. Unmatched ground-truth segments are labeled FN. The F1 score is averaged across trials for each subject:

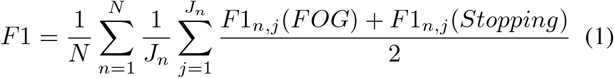

where *N* is the number of subjects, and *J*_*n*_ is the number of trials for subject *n*. For non-FOG/stopping trials with no detected FOG/stopping episodes, an F1 score of 1 was assigned, indicating correct recognition of no FOG/stopping. In the two-step approach, the segmentation block combines FOG and stopping as forward reduction. F1 and F1@50 scores were calculated and averaged per trial as follows:

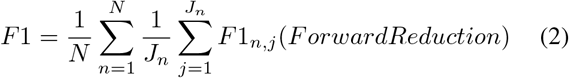

For non-forward reduction trials with no detected episodes, an F1 score of 1 was set, indicating correct detection of no forward reduction. All DL models were trained for 50 epochs with the Amsgrad optimizer, starting at a learning rate of 0.0005, reduced by 5% per epoch, using cross-entropy loss.

In the end-to-end approach, FOG and stopping were treated as positive classes, and F1 and F1@50 scores were computed similarly to the differentiation block using Equation 1.

#### 2) Experiment 2 (Feature comparison for differentiation block)

Next, we we assessed the best model for the differentiation block from Experiment 1, testing it with various modality combinations (IMU, GSR, ECG; IMU+GSR, IMU+ECG, GSR+ECG; IMU+GSR+ECG) to determine the contribution of each modality. We further analyzed the impact of individual features on the model trained with all multimodal data (IMU+GSR+ECG). To ensure an unbiased evaluation, the differentiation block was provided with expert annotations, as illustrated in Figure 5. All experiments were conducted using LOSO-CV, with hyperparameters tuned according to the settings from Experiment 1.

Model performance was assessed using AUROC and F1 scores, calculated as formula 1. Moreover, for feature importance analysis, we used the SHapley Additive exPlanations (SHAP) method [60] to quantify each feature’s impact, which employs Shapley values from cooperative game theory to determine the impact of each feature on the model’s predictions. SHAP decomposes a prediction *f* (*x*) into contributions from each feature *x*_*i*_:

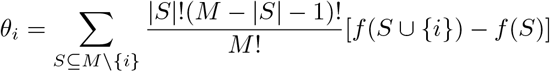

Here, *S* is a subset of features excluding *i*, and *f* (*S*) is the model prediction for subset *S*. The overall prediction is represented as:

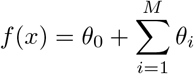

where *θ*_0_ is the average output across the dataset, and *θ*_*i*_ are the SHAP values for feature *i*. A beeswarm plot visualizes these SHAP values, where a SHAP value of -1 indicates a strong impact on predicting FOG, and 1 indicates a strong impact on predicting stopping.

#### 3) Experiment 3 (FOG detection performance comparison)

Finally, we compared both approaches using IMU data alone and all data. In the two-step approach, IMU data was used for segmentation, and the differentiation block was trained on either all features or just IMU features generated from the model-predicted segments. In the end-to-end approach, models were fed with either IMU, GSR, and HR signals or IMU data alone, with HR replacing raw ECG to align sampling rates with IMU and GSR.

All experiments used LOSO-CV, with hyperparameters tuned and training strategies consistent set as in experiments 1 and 2. Model performance was evaluated using F1 scores, calculated using Equation1. FOG severity was assessed from a clinical perspective using the percentage of time frozen (%TF) and the number of FOG episodes (#FOG). Consistency between model predictions and expert annotations was measured with the intra-class correlation coefficient (ICC(2,1)).

### E. Statistics

We used Repeated Measures ANOVA to test for significant differences in F1 scores between models. Pairwise comparisons were performed with paired Student’s t-tests (N = 18, corresponding to the number of subjects), adjusted for multiple comparisons using the Li correction [61]. Levene’s and Shapiro-Wilk tests assessed variance homogeneity and normality. All analyses were conducted at a 0.05 significance level, using Python libraries (SciPy 1.7.11, statsmodels 0.13.2, pingouin 0.3.12) and the R package scmamp 0.2.55.

## IV. Results

### A. Model selection

In the first experiment, we aimed to conduct a model selection study for both the two-step approach and the end-to-end approach to ensure that subsequent experiments were evaluated using the best-performing models.

We began by evaluating models for both blocks in the two-step approach, namely the segmentation block and the differentiation block. As shown in Table IV, ASFormer emerged as the top-performing model for detecting forward movement reduction based on IMU signals, making it the preferred choice for the segmentation block. For the differentiation block, XGBoost achieved the highest F1, F1@50, and AUROC scores for classifying forward movement reduction segments, as indicated in Table V, leading to its selection for this block.

Next, we compared models for the end-to-end approach. Table VI indicates that MS-TCN excelled in detecting FOG and stopping using IMU, GSR, and HR signals, and was therefore selected for the segmentation block.

**TABLE IV.**
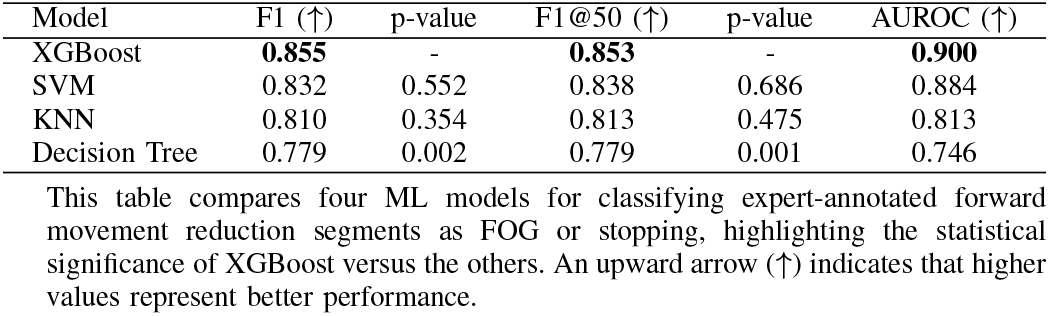
Results of different ML models as differentiation block

**TABLE V.** Results of different DL models as segmentation block

| Model | F1 (↑) | p-value | F1@50 (↑) | p-value |
| --- | --- | --- | --- | --- |
| ASFormer [50] | **0.772** | - | **0.752** | - |
| MS-TCN [49] | 0.771 | 0.930 | **0.752** | 0.965 |
| Dilated TCN [48] | 0.664 | 0.012 | 0.513 | <0.001 |
| Bi-LSTM [47] | 0.548 | <0.001 | 0.472 | <0.001 |

This table compares four DL models for detecting forward reduction segments, with ASFormer showing significant improvement over all models except MS-TCN. An upward arrow (↑) indicates that higher values represent better performance.

**TABLE VI.** Results of different DL models for the end-to-end approach

| Model | F1 (↑) | p-value | F1@50 (↑) | p-value |
| --- | --- | --- | --- | --- |
| MS-TCN [49] | **0.771** | - | **0.759** | - |
| ASFormer [50] | 0.719 | 0.028 | 0.705 | 0.151 |
| Dilated TCN [48] | 0.636 | 0.002 | 0.564 | 0.002 |
| Bi-LSTM [47] | 0.729 | 0.247 | 0.707 | 0.627 |

This table compares four DL models for detecting non-FOG and FOG segments, with p-values indicating the statistical significance of MS-TCN compared to the others. An upward arrow (↑) indicates that higher values represent better performance.

### B. Feature comparison for differentiation block

In the second experiment, we aimed to evaluate the best-performing model for the differentiation block of the two-step approach identified in the first experiment (i.e., XGBoost) with different modality combinations (IMU, GSR, ECG; IMU+GSR, IMU+ECG, GSR+ECG; IMU+GSR+ECG). Table VII shows that the model trained with all IMU+GSR+ECG features (all-modality) achieved the highest F1 and F1@50 and the second-highest AUROC score. It classified FOG episodes and stopping (for all trials in both ST and DT) with the fewest errors, showing significantly higher F1 and F1@50 scores (all p *<* 0.001) compared to models without IMU features (GSR, ECG, GSR + ECG). While the all-modality model also achieved higher F1 scores and AUROC than those with IMU features (IMU, IMU + GSR, IMU + ECG), the differences were not statistically significant. Note that models using only GSR or ECG data performed only marginally better than random guessing, as indicated by AUROC scores just above 0.5, suggesting limited effectiveness in differentiating FOG from stopping episodes on these signals alone. When classification results were separated for ST and DT trials, the all-modality model improved correct identification of FOG episodes in ST trials from 75.98% to 79.90%, with fewer FNs and FPs, but decreased performance in DT trials from 85.45% to 84.55%. This indicates that physiological data improves performance slightly for ST trials but not for DT trials.

**TABLE VII.**
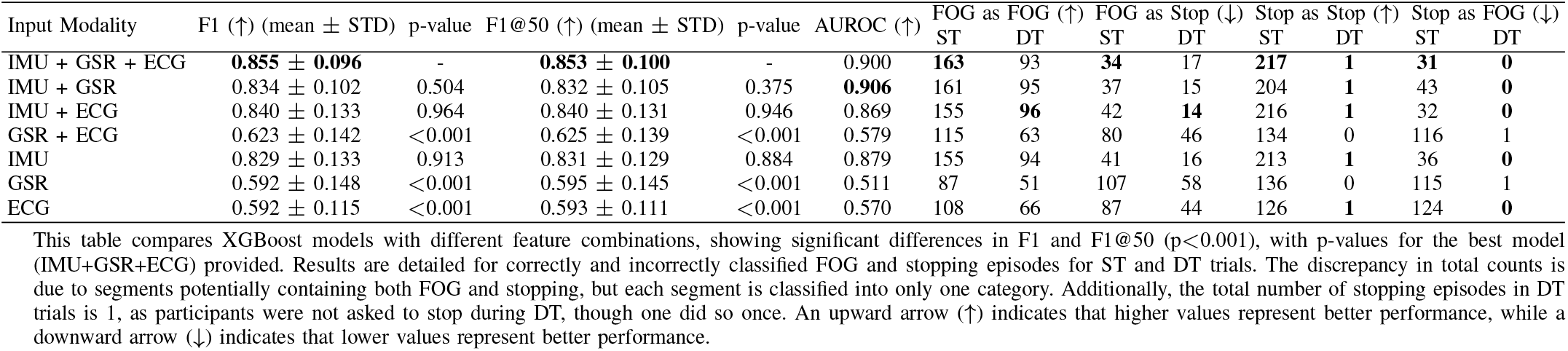
Comparison of the differentiation block (XGBoost) trained using various feature combinations

Feature importance for the all-modality XGBoost model is visualized in Figure 6 through a beeswarm plot, highlighting the top 25 contributing features (calculated based on absolute SHAP values using the SHAP method) in differentiating between FOG and stopping. The plot shows that most influential features are derived from the IMU, with only six from GSR and five from ECG. This indicates that IMU features substantially contribute to the model’s predictive capability, while the contribution of the physiological features is relatively limited. Moreover, the beeswarm plot highlights key feature influences on the model’s predictions. For example, the model is more likely to predict an episode as FOG when it detects a higher freezing index both before and after the episode, as indicated by negative SHAP values. Furthermore, increases in the standard deviation of GSR (C0) and heart rate (HR) from the period before the episode to during the episode also influence the model towards predicting FOG.

**Fig. 6.**
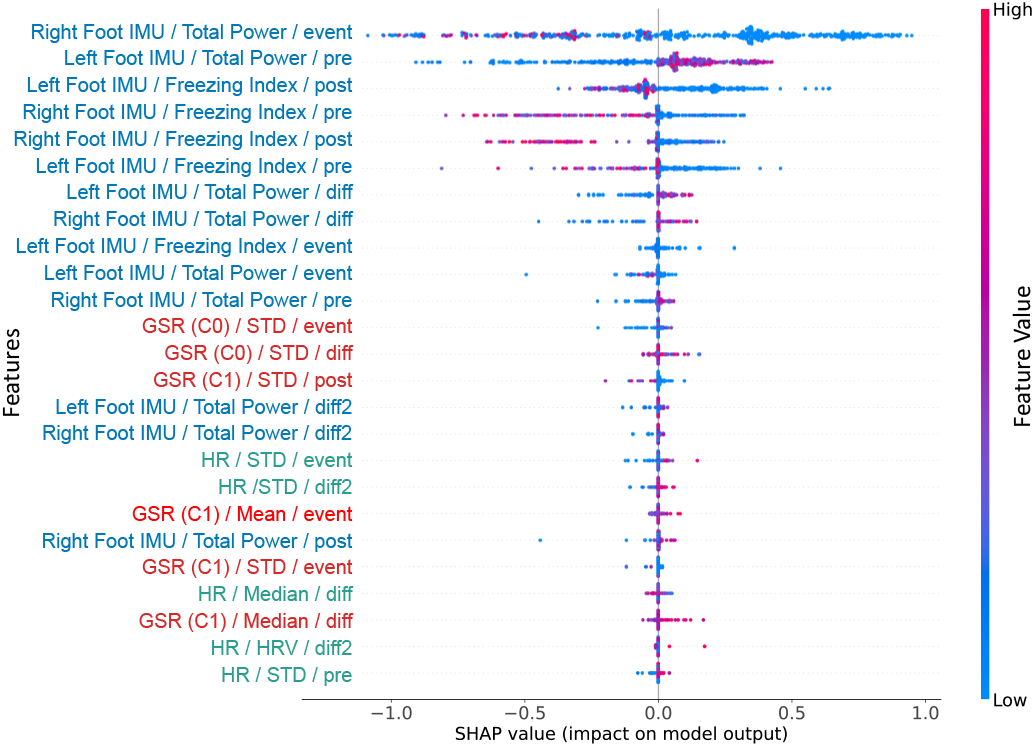
Beeswarm plot of the top 25 contributing features in the XGBoost model. This plot displays SHAP values for each feature, listed vertically, across all input episodes. Points intensify in red with higher feature values and in blue with lower values, highlighting how feature values contribute to the prediction. The naming of the features is organized as follows: Modality / Feature Name / Extracted Window. For the extracted window, “diff” represents the difference between the event and pre-event window, while “diff2” represents the difference between the post-event and event window.

### C. FOG detection performance comparison

Finally, we compared the performance of both the two-step and end-to-end approaches using IMU data alone versus the full multimodal data (IMU+GSR+ECG). In the two-step approach, ASFormer was utilized for the segmentation block, and XGBoost for the differentiation block. For the end-to-end approach, the MS-TCN model was employed to directly detect FOG and stopping events from the multimodal signals.

#### 1) The two-step approach

The two-step approach was evaluated (Table VIII), with the all-modality model achieving an F1 score of 0.728 and an F1@50 score of 0.725, showing high agreement with expert annotations (ICC(%TF) = 0.869, ICC(#FOG) = 0.919). The IMU-only model scored similarly (F1 = 0.731, F1@50 = 0.727) with no significant difference between the two models (p=0.792 and 0.887), indicating a limited impact from including GSR and ECG data.

**TABLE VIII.**
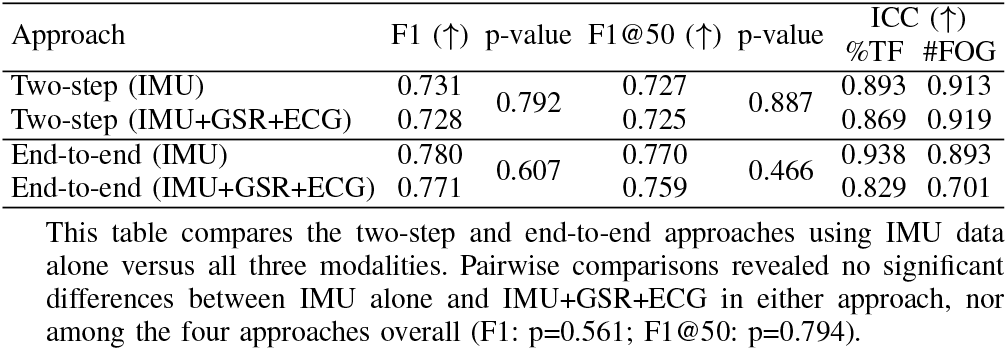
Results of the two-step and end-to-end approach

Optimally, the two-step approach would have the segmentation block accurately detect the exact onset and offset of all forward movement reduction episodes, aligning with expert annotations. This would potentially achieving F1 and F1@50 scores of 0.855 and 0.853 (as shown in Table VII).

#### 2) The end-to-end approach

We also evaluated the end-to-end approach with MS-TCN for segmentation. Table VIII shows that MS-TCN trained with all three modalities achieved an F1 score of 0.771 and an F1@50 of 0.759, slightly lower than the IMU-only model (F1=0.780; F1@50=0.770). These differences were not statistically significant (p=0.607 and 0.466), indicating a marginal decrease in detection performance with the inclusion of physiological signals. Moreover, no significant differences were observed between the two-step and the end-to-end approaches in terms of F1 (p=0.561) and F1@50 (p=0.794). While the end-to-end model shows higher F1 scores than the two-step approach, it is less interpretable, as the two-step approach allows for greater exploration of the features used by the differentiation block.

## V. Discussion

This study investigated FOG detection using multimodal data (IMU, GSR, ECG) to assess the possible benefits of integrating physiological signals with IMU data. Unlike previous research that focused on statistical differences between FOG and stopping events [19], [22], we evaluated the performance of an XGBoost model trained to classify FOG and stopping segments annotated by experts. The model performed best with features from all three data modalities, correctly identifying 81.53% of FOG episodes and 85.16% of stopping episodes, with misclassification rates of 16.24% and 11.72%, respectively. Physiological features improved detection more in ST than DT trials, and SHAP analysis revealed that IMU features were the most influential (14 of the top 25 features). This study also introduced two approaches for FOG detection: a two-step approach using ASFormer for segmentation followed by XGBoost for classification, which offers higher interpretability, and an end-to-end approach using MS-TCN for direct classification, functioning as a black box. However, adding physiological features did not significantly improve performance and even led to slightly lower results in terms of F1 and F1@50 in either approach. This suggests the limited utility of physiological data for improving FOG detection. However, the improvement observed in the differentiation block when using expert-annotated episodes as a perfect segmentation model implies that the benefit of physiological data arises primarily when the onset and offset of reduced forward movement are clearly identified. This indicates that the time windows selected for analyzing physiological features are critical for maximizing their effectiveness in FOG detection.

Previous studies have identified significant differences in physiological features among normal gait, FOG, and stopping episodes using specialized criteria for statistical testing [19], [23], [27], [28]. Our findings suggested limited added value of these signals for FOG detection. We interpret the limited discriminatory effectiveness of physiological data with regard to the following four factors. Firstly, physiological signals change slowly, requiring longer window periods (typically 3 seconds) that can introduce noise from multiple short FOG or stopping episodes. Signals with greater temporal resolution, such as IMU [12] and EEG [37], [38], may be better suited for future studies. Secondly, rapid sequences of movements, such as turning and sitting to standing, introduce motion artifact that complicates distinguishing physiological changes between consecutive FOG episodes [19], [28]. Thirdly, variations in physiological responses to environmental stimuli make uniform responses difficult [28]. Fourthly, cognitive load from DT complicates the differentiation between FOG and stopping using physiological signals, as DT may also influence patients’ stress levels [19], [29], [62]. Finally, the study was conducted in a controlled laboratory setting, ensuring consistent factors such as room temperature and lighting, which is impractical to replicate in real-life scenarios. These limitations suggest that while physiological data provides valuable insights into the mechanisms of FOG, its utility for FOG detection is constrained by inherent variability and signal dynamics.

This study faces several limitations. First, this study used only early fusion for the end-to-end approach. Exploring middle fusion [63] or models that handle different temporal resolutions could better integrate IMU and physiological data, as physiological signals vary more slowly and do not need the 64 Hz sampling rate as for the IMU data. Secondly, the variability in FOG frequency among participants mirrors the range of FOG severity typically seen in clinical settings but could impact the evaluation of physiological data’s usefulness. For participants with fewer FOG episodes, the limited number of data segments might make it harder to assess the contribution of physiological signals. On the other hand, participants with more frequent episodes may generate noisier data due to the close succession of episodes, which could reduce the effectiveness of physiological data in distinguishing FOG from non-FOG events. Thirdly, our study aimed to identify the benefits of physiological data for FOG detection as an initial step toward real-world applications. Although the TUG task with volitional stopping allows for investigating the distinction between stopping and FOG during straight-line walking and turning, it does not consider specific activities related to gait initiation, such as standing up from a chair or turning in place before starting to walk. Our dataset was not designed to capture these transitional movements, which could present valuable opportunities for future multimodal data collection. However, incorporating such scenarios introduces challenges in identifying robust features that are both specific and sensitive enough to function reliably at the individual patient level across diverse conditions. Finally, our dataset (N=18, #FOG=314) is comparable to studies such as Daphnet (N=10, #FOG=237) [40], O’Day *et al*. (N=16, #FOG=211) [10], CuPid (N=18, #FOG=237) [43], Rempark (21 subjects, 1321 episodes) [64], and Zhang *et al*. (N=12, #FOG=334) [38]. However, a larger and more diverse cohort is needed to fully evaluate the impact of physiological data and enhance generalizability.

## VI. Conclusion

Advancing free-living FOG detection requires accurately distinguishing FOG from stopping episodes. This study proposed two approaches for analyzing the possible benefits of physiological data over IMU data for FOG detection. The twostep approach used ASFormer to identify reduced forward movement segments, which were then classified as FOG or stopping by an XGBoost model. The end-to-end approach utilized MS-TCN to directly detect FOG and stopping. Evaluations showed that adding physiological signals had limited benefit on performance in both approaches.

## Data Availability

The datasets analyzed during the current study are not publicly available due to restrictions on sharing subject health information.

